# Heterogeneity in *Mycobacterium tuberculosis* immunoreactivity in young children in Blantyre, Malawi: a community-based survey

**DOI:** 10.64898/2026.05.21.26349011

**Authors:** Hannah M Rickman, Mphatso D Phiri, Hannah Mbale, Helena RA Feasey, Marriott Nliwasa, George Chagaluka, James A Seddon, Henry C Mwandumba, Katherine C Horton, Marc YR Henrion, Tisungane Mwenyenkulu, Kuzani N Mbendera, Emily S Nightingale, Elizabeth L Corbett, Peter MacPherson

**Affiliations:** Clinical Research Department, London School of Hygiene & Tropical Medicine; Malawi Liverpool Wellcome Research Programme; Department of Clinical Sciences, Liverpool School of Tropical Medicine; Helse Nord TB Initiative, Kamuzu University of Health Sciences; School of Medicine, University of St Andrews; Department of Paediatrics, Queen Elizabeth Central Hospital, Blantyre; Department of Infectious Disease, Imperial College London; Desmond Tutu TB Centre, Department of Paediatrics and Child Health, Stellenbosch University; Department of Infectious Disease Epidemiology, London School of Hygiene & Tropical Medicine; Malawi National Tuberculosis and Leprosy Elimination Programme; School of Health and Wellbeing, University of Glasgow

## Abstract

**Background:** As tuberculosis (TB) incidence declines, transmission increasingly concentrates into vulnerable populations. There is an urgent need for affordable surveillance strategies to monitor trends, identify high-risk groups and target interventions. *Mycobacterium tuberculosis* (Mtb) immunoreactivity surveys indirectly detect transmission and therefore undiagnosed infectious disease.

**Methods:** We conducted a cross-sectional community-based interferon-gamma release assay (IGRA) survey in children aged 1-4 years in Blantyre, Malawi. Community-representative participants were recruited using novel convenience sampling in health facilities alongside random household sampling, and tested for Mtb immunoreactivity using QFT-Plus IGRA. We constructed hierarchical Bayesian logistic regression models for IGRA positivity, with neighbourhood-level random effects.

**Findings:** Of 1,545 participants, 102 (6.6%) had a positive IGRA: an annual risk of Mtb infection (ARTI) of 2.7% (95% CrI 2.2-3.2%). Immunoreactivity was higher in the poorest third of households (8.7% vs 4.9%; adjusted odds ratio: 1.88, 95% CrI 1.08-3.01) compared to the richest, but was not associated with HIV exposure, malnutrition or reported household TB exposure. There was substantial between-neighbourhood heterogeneity (ARTI range 1.1-4.1%). There was no association between neighbourhood-level TB case notifications and ARTI.

**Interpretation:** An innovative convenience sampling approach identified a high burden and substantial spatial variation of recent TB transmission, which did not correspond to case notification rates. This strategy could support identification of high-risk populations, monitoring of trends and targeted public health interventions.

**Funding:** This work was funded by the Wellcome Trust [225482/Z/22/Z, 200901/Z/16/Z, 206575/Z/17/Z], and by UK aid from the UK government [to MDP, KCH, MYRH, ELC, PM; “Leaving no-one behind: transforming gendered pathways to health for TB”; 2018/S 196-443482]; however the views expressed do not necessarily reflect the UK government’s official policies.

**Research in context:** *Evidence before this study:* Tuberculin skin test surveys were widely used in the 20^th^ century to estimate population-level tuberculosis (TB) burden, particularly amongst school-aged children, but are limited by cross-reactivity with Bacillus Calmette-Guerin vaccination and non-tuberculous mycobacteria. For a previous meta-analysis we searched Medline Embase, Global Health databases, Science Citation Index Expanded, and Global Index Medicus, using terms related to *Mycobacterium tuberculosis* (Mtb) immunoreactivity and epidemiological study type (1/1/1993 to 31/12/2022; no language restrictions); we performed an updated search on 10/3/2026 restricted to interferon-gamma release assay (IGRA) surveys. Overall, we found four IGRA surveys which included results from children under five. None were designed to assess Mtb immunoreactivity as a measure of recent population-level transmission, and none reported neighbourhood-level data. From Malawi, we identified no community-representative Mtb immunoreactivity surveys from the past decade, and none using IGRA.

*Added value of this study:* We conducted a cross-sectional, community-representative IGRA survey among children under 5 years in Blantyre, Malawi, using pragmatic and scalable methodologies based on convenience clinic sampling and community satellite imagery. Because immunoreactivity in this age group reflects recent exposure, our study provides a proxy measure of ongoing transmission, and therefore of infectious TB disease within the community. We found a substantial burden of Mtb immunoreactivity in young children, and explore the individual- and neighbourhood-level risk factors associated with TB transmission, highlighting priority neighbourhoods for targeted interventions.

*Implications of all the available evidence:* Mtb immunoreactivity surveys have historically provided important insights into TB epidemiology, particularly in settings with falling incidence. Our findings suggest that a pragmatic methodology based on IGRA testing of young children may allow monitoring of trends and identification of high-risk groups and neighbourhoods, supporting targeted interventions in the pursuit of TB elimination.

## Introduction

Tuberculosis (TB) remains the greatest global cause of mortality from any single infectious disease, killing an estimated 1.23 million people in 2024[1]. As global prevalence falls, TB transmission, maintained by undiagnosed disease, becomes increasingly concentrated into vulnerable populations and geographies[2,3], and public health interventions need increasingly efficient targeting to avoid low yield and high costs[4]. Surveillance based on *Mycobacterium tuberculosis* (Mtb) immunoreactivity, indicating previous exposure and infection with Mtb, was widely used by countries with declining TB epidemics in the past century, and may add valuable information to current surveillance approaches[5].

Unlike for many other pathogens, antibodies are not a reliable indicator of Mtb exposure, with immunosurveillance instead requiring evidence of T-cell reactivity using tuberculin skin tests (TSTs), interferon-gamma release assays (IGRAs) or novel skin tests. Mtb immunoreactivity in young children, by definition, provides evidence of *recent* infection (within their lifetimes), implying ongoing Mtb transmission from untreated infectious TB in the community[5]. Young children are at high risk of progression to severe disease and at lower risk of side effects from TB preventive treatment (TPT), and may therefore be more likely to benefit from TPT than other groups[6,7].

Mtb immunoreactivity surveys offer complementarity to disease surveillance methodologies, such as programmatic monitoring of case notification rates (CNRs) or TB disease prevalence surveys. CNRs only capture individuals diagnosed with TB through healthcare systems, thereby excluding the crucial “missing millions” – the estimated quarter of people with TB who go undiagnosed annually[1]. The World Health Organization (WHO) has previously recommended TB disease prevalence surveys in high-prevalence countries with likely low case-detection rates[8], but these require extremely large sample sizes, are usually only powered for national-level precision, and are prohibitively expensive in constrained healthcare systems[5].

Malawi, in Southern Africa, has reported remarkable progress towards TB elimination[1,9,10]. Blantyre, Malawi’s second largest city, is a densely-populated, resource-limited urban setting with an estimated adult HIV prevalence of 15%[11], which exemplifies many of the TB surveillance challenges discussed above. Blantyre’s TB prevalence has declined substantially from 1,014 per 100,000 adults in 2014 to 215 per 100,000 in 2019-2020, likely driven by strengthened decentralised TB services, successful HIV care scale-up, and intensive community-wide TB case-finding[9]. TB burden is unevenly distributed, with genomic analysis suggesting geographic areas of higher transmission[12]. TB CNRs vary significantly across Blantyre[13], but show evidence of an “inverse care law”: notifications are lower in poorer communities with worse healthcare access[14], suggesting that prioritising apparently “high-burden” areas with high CNRs may in fact divert resources away from areas of underdiagnosis. In this epidemiological context, cross-sectional disease prevalence surveys are very expensive and may offer limited additional insights: a 2019-2020 TB disease survey enrolled over 15,000 adult Blantyre residents and diagnosed just 29 individuals with bacteriologically-confirmed TB[9], generating limited power to identify high-burden areas[13].

As TB epidemics decline, efficient new approaches are required to reliably identify areas of higher recent TB transmission and guide targeted interventions. In a time of critical funding shortages, it is essential that these are pragmatic and affordable. We therefore performed a Mtb immunoreactivity survey utilising an innovative clinic-based convenience-sampling approach in children aged 1-4 years in high-risk areas of Blantyre, to determine the annual risk of Mtb infection and identify high-risk individuals and neighbourhoods.

## Methods

Detailed methods are in the published study protocol[15].

### Setting

Participants were recruited November 2022-March 2024 from 33 informal urban neighbourhoods of Blantyre, Malawi. Neighbourhoods were based on Community Health Worker catchment areas, encompassing the approximate catchment areas of the three primary health clinics (PHCs) in Blantyre with the highest number of TB case notifications[15].

### Participants & recruitment

Recruitment followed two approaches: clinic-based convenience sampling in the three PHCs, and random household sampling in the study neighbourhoods. We included children aged 12-60 months normally resident in the study neighbourhoods. There were few exclusion criteria (see below), aiming to recruit a community-representative sample.

### Clinic convenience sample

In the clinic convenience sampling approach, eligible children were identified from those accessing or accompanying others to routine primary health services (e.g. vaccination clinics, check-ups, antenatal care, family planning). Children attending for their own symptomatic illness, or for TB/HIV care, were ineligible. Caregivers waiting for services were approached sequentially until daily recruitment targets were met.

### Random household sampling

Enumeration of possible household structures was performed using an open-access dataset of building footprints from OpenBuildings[16]. Population-weighted random samples were drawn in each neighbourhood, and selected locations visited by study teams, with all age-eligible children in the household approached for recruitment.

### Sample size

Sample sizes were based on precisely estimating an immunoreactivity prevalence of 5% with absolute precision of ±1% and 95% confidence, requiring 1,825 children 1-4 years with a QFT-Plus result, or 100 children with a positive result.

### Laboratory methods

A 4mL venous blood sample was taken for IGRA testing using QuantiFERON-TB Gold Plus (QFT-Plus, Qiagen).

### Results & follow-up

Where possible, we repeated QFT-Plus for participants with indeterminate results. Participants with a positive IGRA were clinically reviewed and referred to the paediatric TB clinic if symptomatic, or to local TB clinics for consideration of TPT if asymptomatic, following Malawi National Guidelines. Their household contacts were offered TB disease screening.

### Covariables

Covariables including HIV status, maternal HIV, Bacillus Calmette-Guérin (BCG) vaccination, previous TB treatment and TB household exposure were ascertained through caregivers’ report and patient-held health records. Wealth status was determined using a proxy means test developed from the Malawi Integrated Household Survey[17,18]. Mid-upper-arm circumference (MUAC) was measured and an age- and sex-adjusted z-score calculated, with malnutrition defined as a z-score ≤-2. Home locations were captured using direct GPS capture for household recruits; for clinic participants, a validated electronic participant location (ePal) app was used to assist participants to geolocate their homes using satellite images and local landmarks[14].

Neighbourhood-level poverty estimates were extracted from a 2019-2020 TB/HIV prevalence survey[9]. Neighbourhood-level HIV prevalence estimates were previously generated using spatially-explicit Bayesian regression from various data sources[11]. Enhanced surveillance of TB case notifications has been performed in Blantyre since 2011; neighbourhood-level CNRs were calculated for January 2021-June 2023[14]. Denominators used 2020 unconstrained gridded population estimates from WorldPop, which were also used to calculate neighbourhood population density[19].

The outcome was a positive QFT-Plus IGRA; indeterminate results were excluded from prevalence calculations. We estimated annual risk of Mtb/TB infection (referred to with the commonly-used acronym ARTI) using the formula ARTI= 1-(1-prevalence)^1/mean age^ and calculated 95% credible intervals by jointly modelling prevalence and mean age as probabilistic quantities using a beta posterior distribution and normal distribution respectively, then drawing 10,000 samples from the joint posterior distributions and calculating the corresponding infection risk to develop a posterior distribution of ARTI estimates. As the estimated risk difference between clinic and household recruits was negligible, we pooled these groups for subsequent analyses.

### Data analysis

Analyses used R version 4·2 and the ‘brms’ interface to Stan. We constructed hierarchical Bayesian logistic regression models for an outcome of positive IGRA, with neighbourhood-level random effects, and calculated odds ratios by summarising 8000 posterior draws. We then explored the addition of individual- and neighbourhood-level covariables to the model, based on i) evidence of effect from univariable analyses and ii) *a priori* knowledge of TB transmission predictors. Models were fit with weakly-informative priors and 4 chains with 5000 iterations. Model convergence was checked by inspection of trace plots, effective sample sizes and 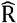 values, with posterior predictive checks used to assess model fit. We assessed collinearity by inspecting a correlation matrix and measuring variance inflation factors, including only non-colinear variables.

We constructed a first model for absolute Mtb immunoreactivity risk associated with residence in a specific neighbourhood, by adjusting for age and sex only and summarising neighbourhood-level random effects (expressed as odds ratios [ORs]). We then added additional individual- and neighbourhood-level covariables to measure their association with Mtb immunoreactivity, and subsequently constructed a second adjusted model, incorporating individual- and neighbourhood-level covariables identified as associated with the outcome, summarising neighbourhood-level random effects again to understand the extent of residual neighbourhood-level variation unaccounted for by the given covariables. Moran’s I was used to assess for autocorrelation in model residuals, using an adjacency matrix based on shared neighbourhood boundaries.

We predicted neighbourhood-level ARTI from the posterior distributions of the age- and sex-adjusted model. We hypothesised a scenario in which a TB programme targeted the highest-risk quartile of neighbourhoods, and therefore identified eight “priority” neighbourhoods with the highest modelled ARTI, and compared those to the neighbourhoods with the highest CNRs.

The code to reproduce analyses is available at https://github.com/hannahrickman/ along with a summary dataset (home location has been removed to preserve confidentiality).

### Ethics & informed consent

Caregivers of eligible children were provided with oral and written information about the study and invited to provide individual informed consent in writing or, if illiterate, by a thumbprint observed and countersigned by an independent witness. Ethical approval was received from the Kamuzu University of Health Sciences (Ref P.04/22/3611) and the London School of Hygiene and Tropical Medicine (Ref 2774).

## Results

### Participant characteristics

We screened 2,882 children, of whom 2,581 (90%) were eligible for inclusion and 1,851 (55%) consented for enrolment (Supplementary data 1). IGRA results were available for 1,738 (94%) participants, of which 190 (11%) were indeterminate and excluded from this analysis. In total, we included results from 1,545 participants, with a median age of 2.3 (interquartile range [IQR] 1.5-3.5) years (Table 1). 754 (49%) were female. 297 (19%) had been exposed to maternal HIV, of whom 17 (1.1% overall) were HIV positive. Almost all (1,516, 98%) had received BCG vaccination, and three (0.2%) were reported to have been previously treated for TB. 56 (3.6%) were reported to have had a TB household exposure.

**Table 1.** Participant characteristics, by recruitment location. TB: tuberculosis. BCG: Bacillus Calmette-Guérin. MUAC-Z: age- and sex-adjusted mid-upper arm circumference Z-score. SD: standard deviations. IQR: interquartile range. ARTI: annual risk of Mtb infection. CI: credible interval. ^1^Euclidean distance of home location to nearest primary health clinic Categorical variables are reported as number and percentage. Continuous variables are reported as a median and interquartile range, with the exception of ARTI, which is reported as an overall percentage estimate with 95% credible estimates calculated from the joint posterior distribution of prevalence and mean age, as described in the methods.

| Characteristic | Overall<br>N = 1,545 | Clinic<br>N = 1,134 | Household<br>N = 411 |
| --- | --- | --- | --- |
| **Individual characteristics** |  |  |  |
| Age, years – median (IQR) | 2.31 (1.50, 3.50) | 2.02 (1.36, 3.18) | 3.14 (2.09, 4.00) |
| Female | 754/1545 (49%) | 549/1134 (48%) | 205/411 (50%) |
| HIV positive | 17/1545 (1.1%) | 14/1134 (1.2%) | 3/411 (0.7%) |
| HIV exposure | 297/1544 (19%) | 238/1134 (21%) | 59/410 (14%) |
| Current cough | 253/1545 (16%) | 191/1134 (17%) | 62/411 (15%) |
| Any current TB symptoms | 374/1545 (24%) | 293/1134 (26%) | 81/411 (20%) |
| Previous BCG vaccination | 1,516/1540 (98%) | 1,119/1131 (99%) | 397/409 (97%) |
| Previous TB treatment | 3/1543 (0.2%) | 1/1134 (<0.1%) | 2/409 (0.5%) |
| **Caregiver-reported general health** |  |  |  |
| Excellent | 461/1545 (30%) | 341/1134 (30%) | 120/411 (29%) |
| Good | 953/1545 (62%) | 695/1134 (61%) | 258/411 (63%) |
| Fair | 115/1545 (7.4%) | 85/1134 (7.5%) | 30/411 (7.3%) |
| Poor | 16/1545 (1.0%) | 13/1134 (1.1%) | 3/411 (0.7%) |
| MUAC-Z below -2SD | 45/1545 (2.9%) | 29/1134 (2.6%) | 16/411 (3.9%) |
| Hospital admission in lifetime | 220/1545 (14%) | 180/1134 (16%) | 40/411 (9.7%) |
| Current school attendance | 418/1545 (27%) | 254/1134 (22%) | 164/411 (40%) |
| Ever travelled outside of Blantyre | 747/1545 (48%) | 537/1134 (47%) | 210/411 (51%) |
| **Household characteristics** |  |  |  |
| Number of household members – median (IQR) | 5.0 (4.0, 6.0) | 4.0 (3.0, 6.0) | 6.0 (4.0, 7.0) |
| **Means score** |  |  |  |
| Poorest tertile | 515/1545 (33%) | 397/1134 (35%) | 118/411 (29%) |
| Middle tertile | 515/1545 (33%) | 415/1134 (37%) | 100/411 (24%) |
| Richest tertile | 515/1545 (33%) | 322/1134 (28%) | 193/411 (47%) |
| **Educational level of household head** |  |  |  |
| Primary or below | 507/1545 (33%) | 364/1134 (32%) | 143/411 (35%) |
| Some secondary education | 377/1545 (24%) | 300/1134 (26%) | 77/411 (19%) |
| Completed secondary or above | 661/1545 (43%) | 470/1134 (41%) | 191/411 (46%) |
| Distance to clinic (km)<sup>1</sup> – median (IQR) | 1.30 (0.77, 2.13) | 1.31 (0.77, 2.09) | 1.27 (0.77, 2.18) |
| TB household contact | 56/1545 (3.6%) | 24/1134 (2.1%) | 32/411 (7.8%) |
| **Outcome** |  |  |  |
| **IGRA result** |  |  |  |
| Negative | 1,443/1545 (93%) | 1,064/1134 (94%) | 379/411 (92%) |
| Positive | 102/1545 (6.6%) | 70/1134 (6.2%) | 32/411 (7.8%) |
| ARTI, % - estimate, 95% CrI | 2.7 (2.2, 3.2) | 2.7 (2.1, 3.4) | 2.7 (1.8, 3.7) |

### Participants by recruitment site

Participants recruited using clinic convenience sampling and random household sampling were similar with respect to sex, HIV status, current cough, caregiver-reported general health, previous TB, and nutritional status (Table 1). However, children recruited from clinics were younger, with a higher prevalence of HIV exposure and current symptoms, and from poorer households. Children recruited from households were more likely to have reported TB household exposure. These differences persisted when adjusted for age (Supplementary data 2). Nevertheless, there was no evidence of difference in ARTI between clinic and household groups, with a predicted ARTI of 2.7% in both (estimated absolute risk difference 0.14%, 95% CrI −1.1 to 1.1%).

### IGRA positivity and ARTI

Overall, 102 (6.6%) participants had a positive IGRA, equating to an ARTI of 2.7% (95% CrI 2.2-3.2%) per year. The estimated prevalence was slightly higher in girls than in boys (7.6% vs 5.7%; adjusted OR [AOR] 1.38, 95%CI: 0.93-2.06) (Table 2). We did not find strong evidence for association between Mtb immunoreactivity and HIV exposure, malnutrition, distance to clinic or caregiver-reported TB household exposure, nor with neighbourhood characteristics such as neighbourhood-level poverty, HIV prevalence or population density. Children living in the poorest third of households were more likely to have Mtb immunoreactivity than the richest tertile (8.7% vs 4.9%; AOR 1.88, 95%CI: 1.08-3.01).

**Table 2.** Risk factors for Mycobacterium tuberculosis immunoreactivity (combined clinic convenience and random household participants). Odds ratios are expressed relative to the reference categories (for categorical variables), or per 1-unit increase in continuous variables. Continuous variables are summarized as median (interquartile range) IGRA: Interferon-gamma release assay. OR: Odds ratio. AOR: Adjusted odds ratio. CrI: Credible interval. MUAC-Z: Age- and sex-adjusted mid-upper arm circumference Z-score. TB: Tuberculosis. ^1^Odds ratios from a multilevel Bayesian logistic regression model, with random intercepts for neighbourhood. ^2^As above, but additionally adjusted for age and sex. ^3^Data missing for one participant. ^4^See methods. ^5^See methods; higher (i.e. less negative) scores represent greater poverty. ^6^CNRs scaled to cases per 1,000 per year (to aid interpretability of odds ratios)

| Characteristic | IGRA Negative<br>n = 1,443 | IGRA Positive<br>n = 102 | OR<sup>1</sup><br>(95% CrI) | AOR<sup>2</sup><br>(95% CrI) |
| --- | --- | --- | --- | --- |
| **Individual variables** |  |  |  |  |
| Age (years) | 2.32<br>(1.48, 3.50) | 2.28<br>(1.70, 3.50) | 1.03<br>(0.87-1.22) | 1.03<br>(0.86-1.22) |
| **Sex** |  |  |  |  |
| Male | 746<br>(52%) | 45<br>(44%) | REF | REF |
| Female | 697<br>(48%) | 57<br>(56%) | 1.38<br>(0.93-2.07) | 1.38<br>(0.93-2.06) |
| **HIV exposure** |  |  |  |  |
| HIV unexposed | 1,164<br>(81%) | 83/101<sup>3</sup><br>(82%) | REF | REF |
| HIV exposed | 279<br>(19%) | 18/101<sup>3</sup><br>(18%) | 0.88<br>(0.49-1.49) | 0.87<br>(0.51-1.43) |
| **MUAC** |  |  |  |  |
| MUAC-Z ≥ -2 | 1399<br>(97%) | 101<br>(99%) | REF | REF |
| MUAC-Z below -2 | 44<br>(3%) | 1<br>(1%) | 0.29<br>(0.03-1.5) | 0.28<br>(0.03-1.43) |
| **Household variables** |  |  |  |  |
| **Household wealth** |  |  |  |  |
| Richest tertile | 490<br>(34%) | 25<br>(25%) | REF | REF |
| Middle tertile | 483<br>(33%) | 32<br>(31%) | 1.29<br>(0.71, 2.14) | 1.34<br>(0.74, 2.21) |
| Poorest tertile | 470<br>(33%) | 45<br>(44%) | 1.87<br>(1.10, 3.03) | 1.88<br>(1.08, 3.01) |
| **Education level of household head** |  |  |  |  |
| Some secondary education | 978<br>(68%) | 60<br>(59%) | REF | REF |
| Primary education or below | 465<br>(32%) | 42<br>(41%) | 1.37<br>(0.91, 2.04) | 1.39<br>(0.91, 2.08) |
| Distance to clinic (km) | 1.30<br>(0.77, 2.12) | 1.32<br>(0.81, 2.25) | 0.98<br>(0.72-1.36) | 0.98<br>(0.72-1.33) |
| **TB household exposure** |  |  |  |  |
| No TB household exposure | 1,391<br>(96%) | 98<br>(96%) | REF | REF |
| TB household exposure | 52<br>(4%) | 4<br>(4%) | 1.05<br>(0.31-2.97) | 1.04<br>(0.32-2.87) |
| **Neighbourhood variables** |  |  |  |  |
| Neighbourhood adult HIV prevalence, %<sup>4</sup> | 17.2<br>(16.6, 18.9) | 17.7<br>(16.6, 20.2) | 1.12<br>(0.96, 1.30) | 1.12<br>(0.95, 1.30) |
| Median poverty score<sup>5</sup> | -5.76<br>(-6.67, -5.11) | -6.05<br>(-6.77, -5.23) | 0.83<br>(0.64-1.07) | 0.82<br>(0.64-1.07) |
| Population density, people per 10,000m<sup>2</sup> | 179<br>(100, 347) | 154<br>(99, 209) | 0.81<br>(0.66, 1.01) | 0.81<br>(0.65, 1.00) |
| TB case notification rate, cases per 1,000 per year<sup>6</sup> | 1.53<br>(1.16, 1.85) | 1.53<br>(1.16, 1.85) | 1.17<br>(0.70-2.08) | 1.17<br>(0.70-2.03) |

### Neighbourhood-level variability

There was substantial between-neighbourhood heterogeneity in Mtb immunoreactivity prevalence in the age- and sex-adjusted hierarchical model, (Supplementary data 5), accounting for around 9% of total variability (intraclass correlation coefficient 0.087, 95% CrI 0.017-0.21). High prevalence of Mtb immunoreactivity was observed in several peri-urban neighbourhoods around Bangwe health centre and in some neighbourhoods around Ndirande (Figure 1A). Predicted neighbourhood ARTI ranged from 1.11% (0.32-2.42%) to 4.11% (1.70-8.30%). Participants in the highest-risk neighbourhood had twice the odds of Mtb immunoreactivity of the overall cohort (OR 1.98, 95% CrI: 0.86-4.57), while those in the lowest-risk neighbourhood had half the odds (OR 0.48, 95% CrI: 0.20-1. 02) (Supplementary data 3).

**Figure 1.**
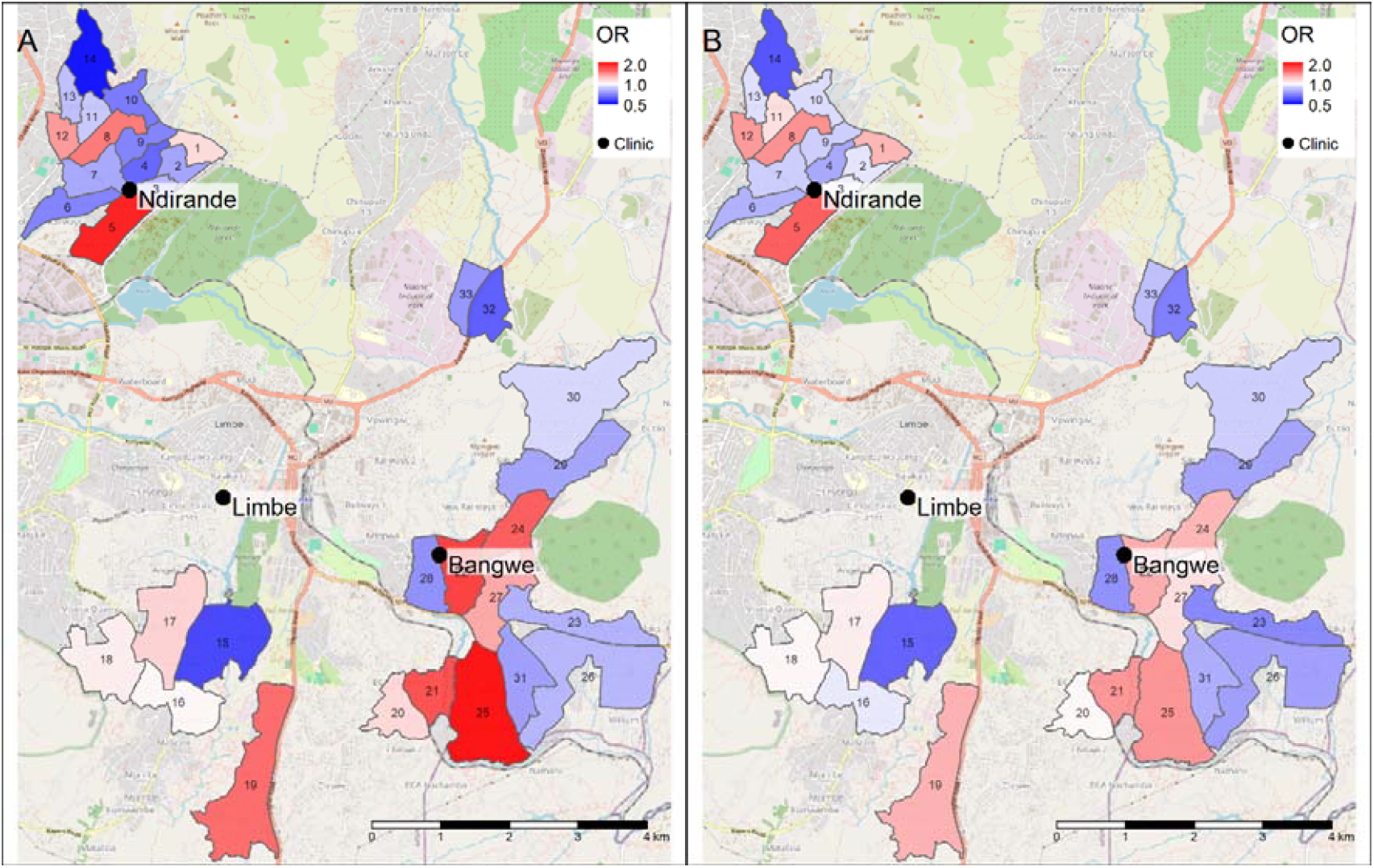
Odds of Mtb immunoreactivity associated with residence in each neighbourhood: neighbourhood-level Mtb immunoreactivity random effects, expressed as odds ratios. A. Age- and sex-adjusted model, showing absolute odds ratios associated with neighbourhoods. B. Model including age, sex, household poverty, neighbourhood HIV prevalence, and population density, showing residual neighbourhood-specific random effects once risk factors are accounted for. Map tile data from OpenStreetMap (https://www.openstreetmap.org/copyright). OR: odds ratios.

Many potentially-relevant covariables also varied spatially (Supplementary data 4); we therefore constructed an extended model which included age, sex, household poverty, and neighbourhood-level HIV prevalence and population density (Supplementary data 5). There was some evidence for higher Mtb immunoreactivity in areas with high HIV prevalence (AOR 1.08 per 1% increase in HIV prevalence; 95% CrI 0.93-1.25)) and with lower population density (AOR 0.84 per person per 10,000m^2^ increase, 95% CrI 0.67-1.04). Adding covariables did not substantially improve model fit (Supplementary data 5), and resulted in a modest reduction in between-neighbourhood variability (ICC 0.055, 95% CrI 0.001-0.179) and the magnitude of the remaining neighbourhood-level random effects (Figure 1B). There was no evidence of residual spatial autocorrelation in either model using Moran’s I (Supplementary data 5).

### Comparison with case notifications

There was no association between areas with high TB CNRs, and high risk of Mtb immunoreactivity (AOR 1.17 per 1 per 1,000 increase in CNR, 95% CrI 0.70-2.03; Figure 2). Of eight hypothetical “priority” neighbourhoods identified as having the highest ARTI, only two were also within the eight areas with the highest CNR.

**Figure 2.**
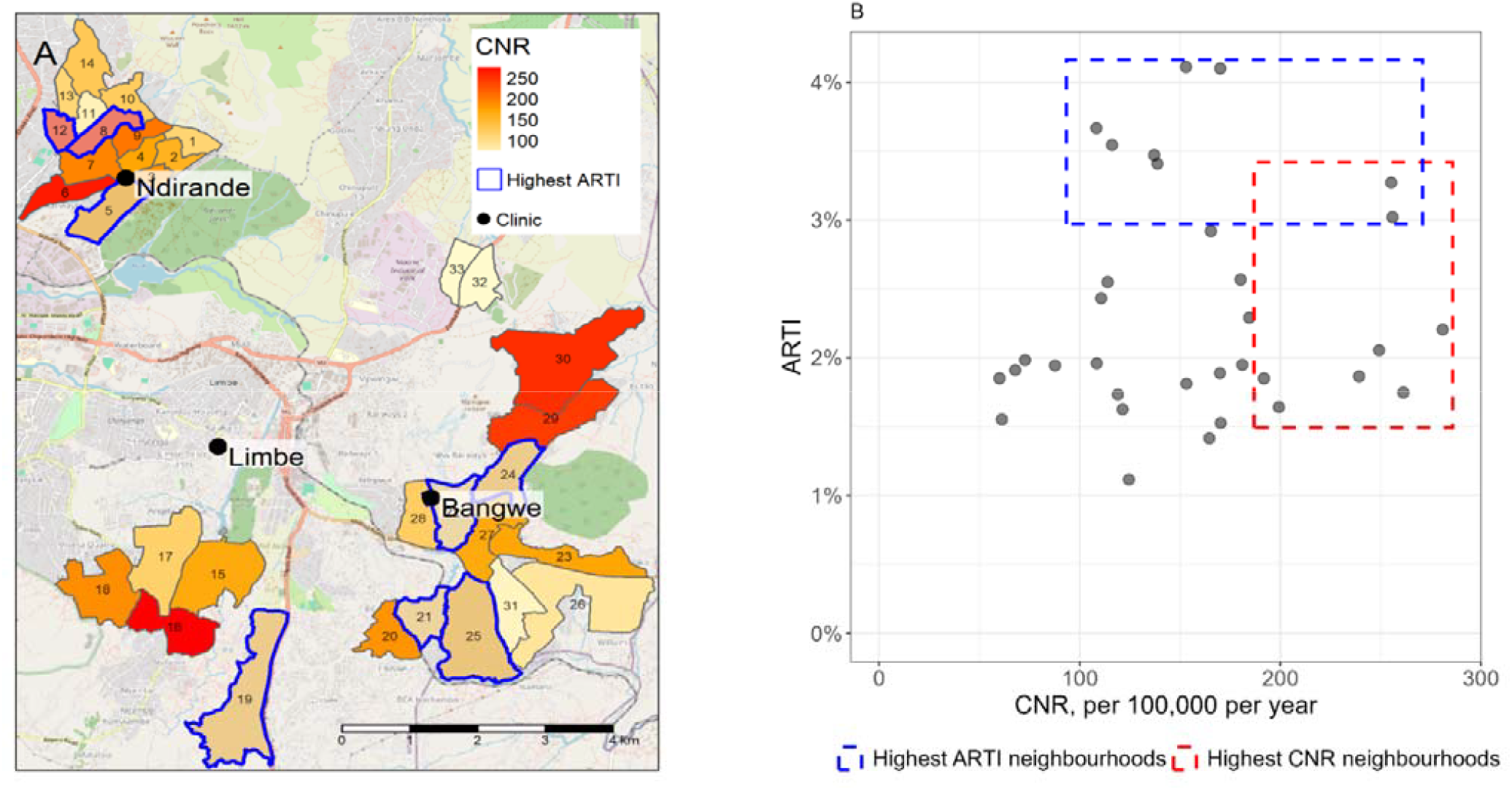
Correlation between neighbourhood-level case notification rates (CNR) and annual risk of Mtb infection (ARTI). Map tile data from OpenStreetMap (https://www.openstreetmap.org/copyright) A. Case notification rates per 100,000 per year, by neighbourhood. Areas highlighted in blue are the eight neighbourhoods with the highest ARTI, which might theoretically be targeted for intervention by a TB programme if they were to prioritise based on ARTI, rather than CNR. B. Neighbourhood-level correlation between CNR and ARTI. Again, the neighbourhoods which would be targeted for intervention based on ARTI are highlighted in blue, while those which would be targeted for intervention based on CNR are highlighted in red.

## Discussion

We used innovative convenience- and random household-sampling methodologies to conduct an Mtb immunoreactivity survey in a city with a rapidly evolving TB epidemic, and found evidence of a substantial (2.7%) annual risk of TB transmission to young children. Mtb immunoreactivity was higher in children from the poorest households and in some peri-urban neighbourhoods, indicating local heterogeneity in TB epidemiology. Our findings suggest that Mtb immunoreactivity surveillance may allow modelling of local TB transmission trends and identification of high-risk neighbourhoods for public health interventions, as well as identifying TB-exposed children who may benefit from TPT.

Global health funding cuts demand that any surveillance methodology optimises cost-effectiveness and efficiency, for example by integrating within existing systems such as primary care. Our clinic-based approach built on a strategy of opportunistically screening clinic attendees for malaria parasitaemia in order to identify high-transmission villages in Chikwawa, Malawi[20]. We did note some differences in the characteristics of participants recruited via clinic-based convenience versus cross-sectional random household sampling. These differences may reflect information bias (data was gathered on household recruits within their own household, and typically from the head of household, which may have affected responses to questions on poverty and maternal HIV status) or true selection differences (children attending clinic for vaccinations or accompanying family members were on average younger), or a combination. Nevertheless, the overall ARTI estimates were very similar, suggesting that in this context clinic-based convenience sampling could offer a cost-effective alternative to random household sampling. Clinic-based surveillance may facilitate linkage to care for participants eligible for TPT and offers opportunities to integrate within multi-disease surveillance systems.

The main individual risk factor for Mtb immunoreactivity was household poverty, with children in the poorest third of households having 1.9-fold higher odds of IGRA positivity compared to the richest tertile. There are multiple interacting mechanisms by which household poverty can increase TB risk, including: crowding, housing conditions, exposure to congregate spaces, nutrition, air quality, healthcare access, and co-association with factors such as HIV and smoking. The relationship between poverty and TB is complex and often context-specific[21,22], but our findings support poverty alleviation and social protection for vulnerable communities as essential components of global efforts to end TB.

We found no association between maternal HIV status and child Mtb immunoreactivity; previous research is mixed[23–25]. Similarly, while malnutrition is strongly associated with TB *disease*, we did not observe an association with Mtb immunoreactivity, in line with the theory that malnutrition may predominantly impact progression from infection to disease[26].

The lack of association between TB household exposure and Mtb immunoreactivity may partially reflect challenges in accurately ascertaining TB exposure, but is compatible with knowledge that most TB transmission to young children occurs outside the household[27]. This supports our hypothesis that, while young children’s TB exposure may not fully reflect community transmission, it is likely to be impacted by their wider local environment. Indeed, as young children are typically less mobile within cities than adolescents and adults, Mtb immunoreactivity in this age group may serve as a powerful sentinel of undiagnosed infectious TB in their neighbourhood.

We found a four-fold variation in estimated ARTI by neighbourhood, indicating possible priority areas for public health action. This is despite the study being conducted within a single, relatively homogeneous city, in purposively selected informal neighbourhoods with higher TB CNRs; transmission is likely even more heterogeneous in more unequal settings. Modelling work suggests that spatial targeting of TB interventions such as active case-finding may disproportionately impact transmission[28]; however very few studies describe *implementation* of spatially-targeted TB interventions in high-incidence settings, in part due to the challenges in identifying high-risk neighbourhoods and in evaluating the impact of such interventions[2,29]. Spatially-concentrated transmission may arise due to a combination of environmental factors such as crowding, distribution of population factors such as HIV prevalence, and local “outbreaks” of transmission[12]. In this study, known risk factors such as HIV prevalence and population density accounted for only a minority of the neighbourhood-level variation in risk, suggesting an additional contribution from local transmission. Furthermore, these relationships were complex. For example, we found weak evidence of a *negative* association between population density and Mtb immunoreactivity: plausible in the Blantyre context, as neighbourhoods around one clinic (Ndirande) are dense but well-established, with relatively good access to public healthcare, whereas lower-density peri-urban regions around a second clinic (Bangwe) represent more recent informal urban expansion. This reinforces the complexities in predicting local TB risk and the need for context-specific epidemiological data.

Importantly from a programmatic perspective, CNRs did not predict Mtb immunoreactivity, highlighting the limitations of CNRs as indicators of undiagnosed infectious disease within communities; indeed, higher CNRs may simply reflect greater access to healthcare[14]. Several TB programmes are targeting high-CNR areas for enhanced intervention, but in this setting only two of the “priority” highest-risk quartile of neighourhoods based on CNR were in the highest quartile of risk measured by ARTI. Combining Mtb immunoreactivity measurements with disease prevalence surveys (in specific settings), genomic sequencing, markers of TB severity, or predictors of underdiagnosis, may add further triangulation and power to understand which areas truly have the highest burden of undiagnosed disease[5,12,13]. Further value in Mtb immunoreactivity surveillance may come in measuring trends rather than in one-off surveys, and future surveys may add further data to this picture.

The 2.7% ARTI was higher than historically observed in limited previous data from Malawi. The 1994 Malawi National Tuberculin survey in 6-11-year-olds estimated an ARTI of 1% (varying from 0.7%- 2.2% by region)[30], while a 2012 TST survey in 2-4-year-olds in rural Northern Malawi, which used a strict 15mm cut-off, found an ARTI of 0.3%[24]. Blantyre has a high TB incidence relative to the rest of Malawi, and the ARTI in our study may have been inflated by purposive selection of urban areas with high case-notifications. Additionally the children recruited lived through the COVID-19 pandemic, which may have caused delays in TB diagnosis and increased community transmission [31]. Furthermore, surveys in older age-groups which estimate ARTI without accounting for reversion may significantly under-estimate true transmission[5,32].

This study has several limitations. Firstly, most participants were recruited through a primary care-based convenience sampling approach. This was partly justified by the high uptake of primary healthcare in Malawi, and the logistical and cost advantages; it is noteworthy that conventional household-based sampling methods are also subject to selection biases. Our approach is supported by similar results from clinic and household-based groups; however, we did note some differences in the characteristics of these children which may suggest the clinic-based group may not have been fully representative. From a statistical perspective, the non-probabilistic sampling approach limits formal inference to the wider population. The utility of convenience sampling is likely to vary between settings: Malawi has an established unified public primary healthcare system, but clinic-based sampling would be more problematic in places with multiple public and private providers, or with more uneven access to care. Broader application would require context-specific validation, such as the hybrid strategy employed here, which uses a smaller, random household-based sample to validate the more pragmatic clinic-based estimates. Integrating Mtb immunoreactivity testing into ongoing programmes such as Demographic Health Surveys or Multiple Indicator Cluster Surveys might improve comparability of results between settings.

There is no “gold-standard” test for Mtb infection, and Mtb immunoreactivity does not fully capture exposure or individual risk. IGRA performance in young children appears comparable to older age groups[33], but there is less data from children under two years. Some of the covariables in our model are themselves estimates (for example HIV prevalence and Worldpop estimates); nevertheless, these reflect the available data sources for policy-makers in settings such as Blantyre.

## Conclusion

TB remains a major global killer, and countries are not on track to achieve ambitious 2030 End TB goals[1]. Context-specific surveillance systems and timely data are required to design targeted strategies to address complex, heterogeneous and changing TB epidemiology. This study highlights the potential of IGRA surveillance using an innovative clinic-based convenience sampling as a tool for monitoring Mtb transmission, identifying high-risk areas, and targeting interventions to optimise use of increasingly limited resources. Mtb infection was associated with poverty, and it is critical to address the social determinants of TB to prevent morbidity and mortality, and to achieve TB elimination goals.

## Supporting information

STROBE checklist

Ethics approval

Ethics approval 2

## Data Availability

The code to reproduce analyses will be made available at https://github.com/hannahrickman/ along with a summary dataset (home location has been removed to preserve confidentiality).

## Supplementary data

**Supplementary data 1.**
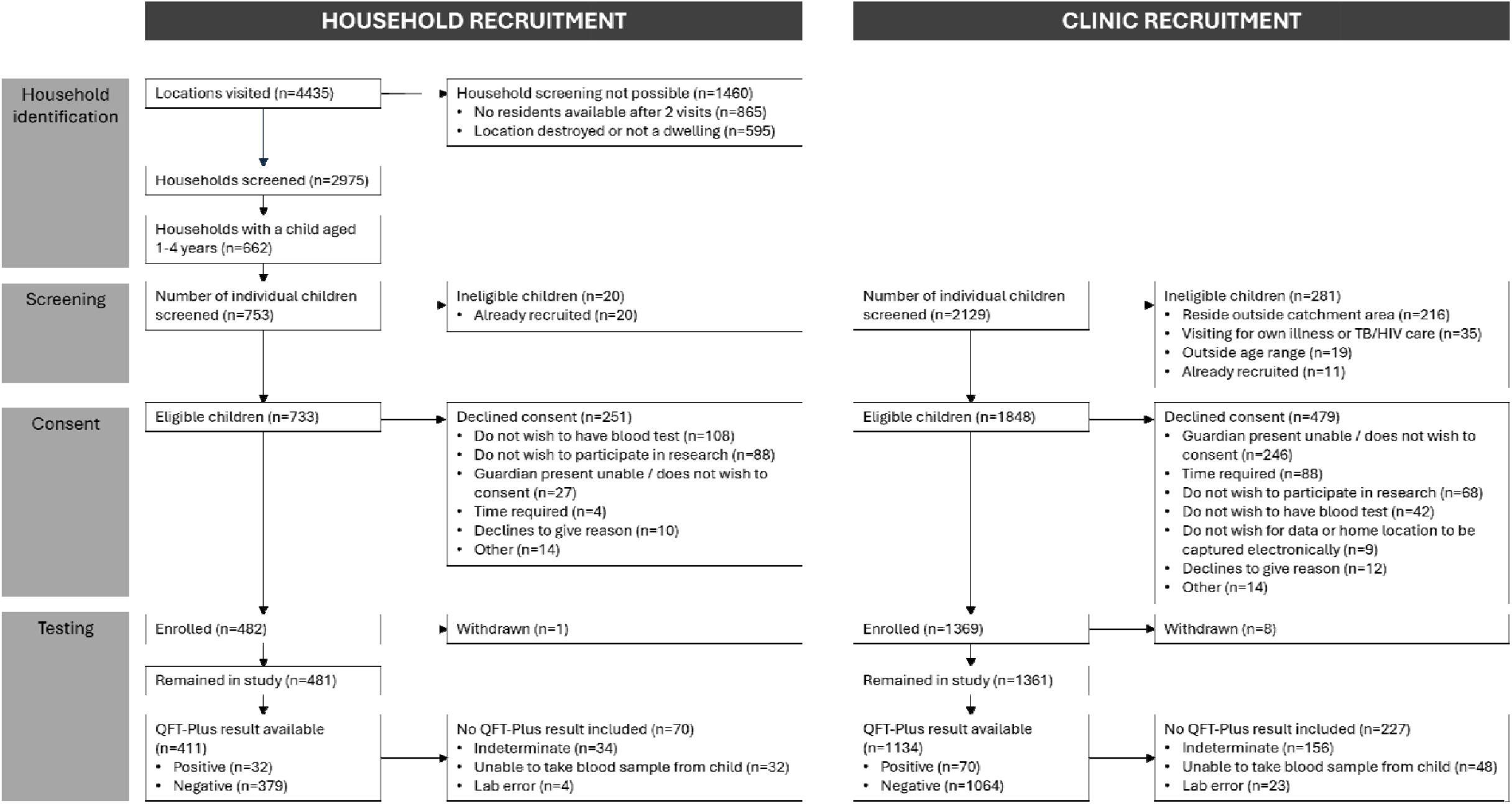
Flow chart of participants screened and recruited into the study.

**Supplementary data 2.**
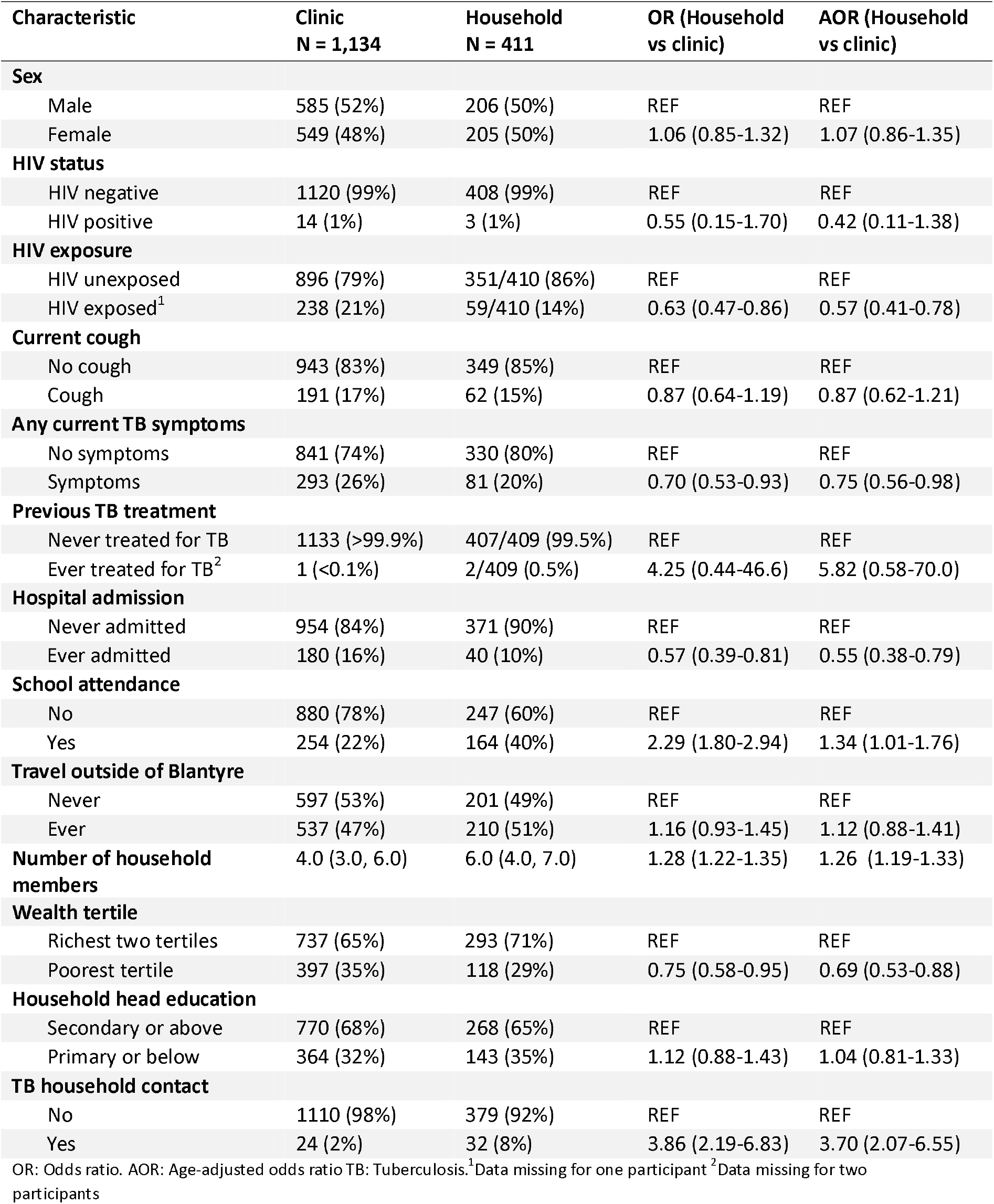
Age-adjusted associations between covariables and recruitment location. Odds ratios and age-adjusted ratios for each characteristic in household, as compared to community, participants, +/- adjusted for age. Odds ratios are expressed relative to the reference categories (for categorical variables), or per 1-unit increase in continuous variables. Continuous variables are summarized as median (interquartile range).

**Supplementary data 3.**
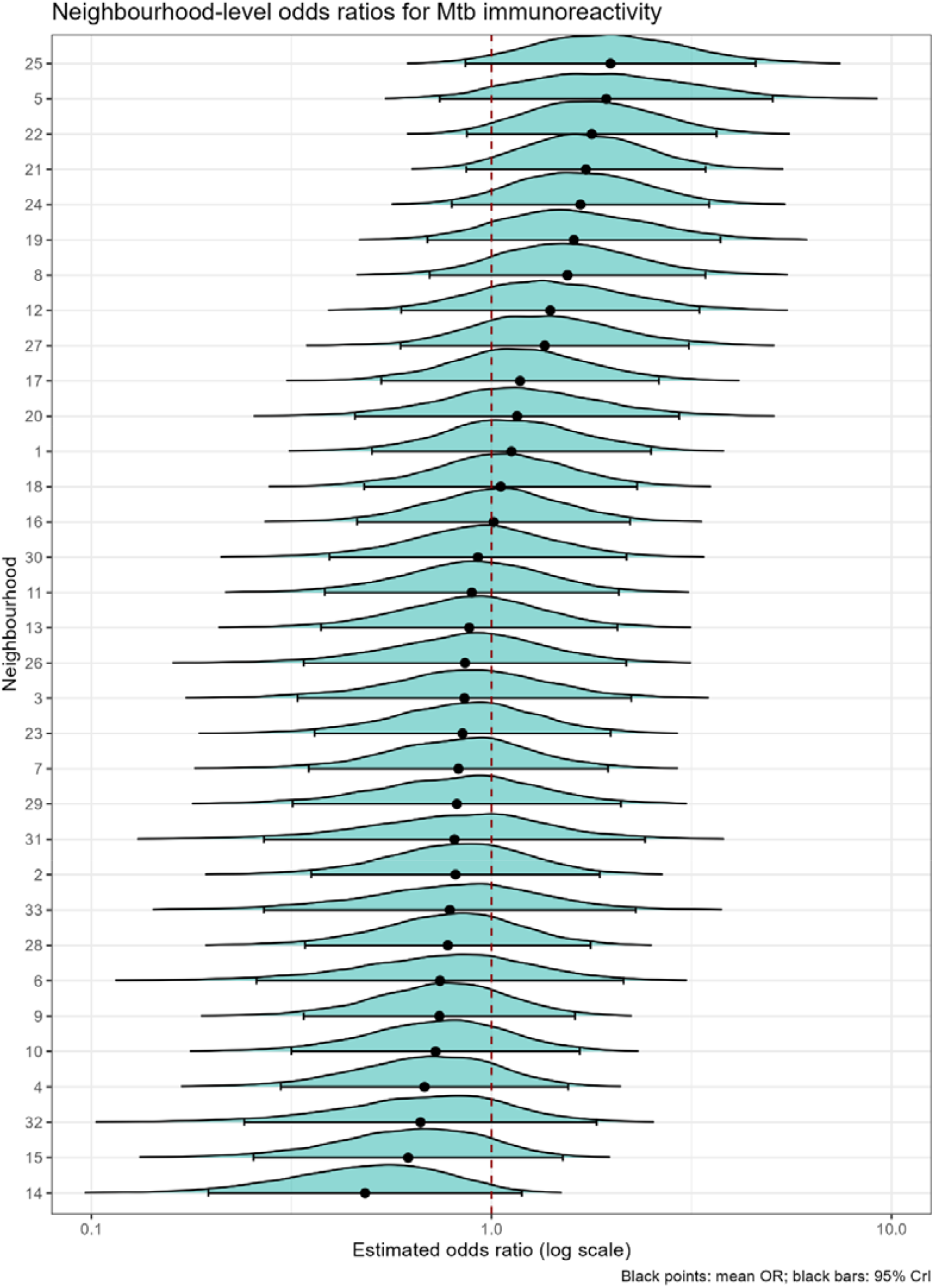
Neighbourhood-level odds for Mtb immunoreactivity. Forest plot showing neighbourhood-level estimated odds ratios (ORs) for Mtb immunoreactivity, relative to a reference neighbourhood with a mean random effect (i.e. OR = 1). Each density ridge represents the full posterior distribution of the neighbourhood-level random effect.

**Supplementary Data 4.**
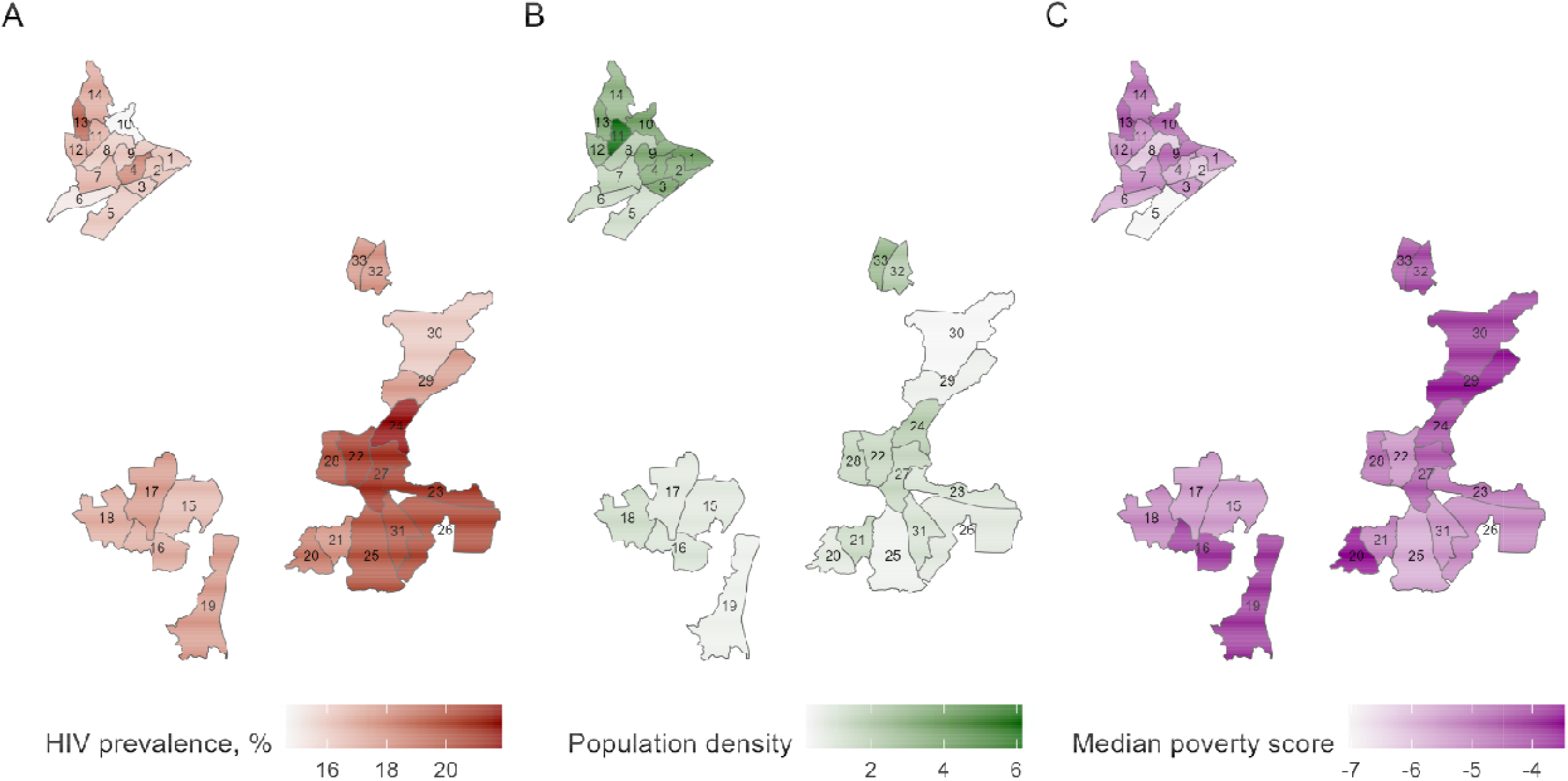
Spatial variation of neighbourhood-level covariables. A. HIV prevalence amongst adults aged 15-49 years (data previously generated using spatially-explicit Bayesian regression from the 2019 survey and two further national HIV prevalence surveys [11]) B. Population density (people per 100m i.e. 10m x 10m), from unconstrained gridded population estimates for 2020 from Worldpop [19] C. Median poverty score using a proxy means test; higher (i.e. less negative) scores indicate higher chance of poverty. Data from a 2019 TB prevalence survey [9].

### Supplementary data 5 - Simple and full models for risk of Mtb immunoreactivity

Baseline model 1:

logit(Pr(QFTP_i_)) = β_0_ + β_1_ ⍰Age_i_ + β_2_ ⍰Sex_i_ + u_neighbourhood[i]_

Extended model 2:

logit(Pr(QFTP_i_)) = β_0_ + β_1_⍰ Age_i_ + β_2_ ⍰Sex_i_ + β_3_⍰ Poverty_i_ + β4 ⍰HIVPrev_neighbourhood[i]_ +β5 ⍰Density_neighbourhood[i]_ + u_neighbourhood[i]_

Where:

QFTP_i_ Binary outcome for individual i (log-odds of QFTP positivity)

β_0_ Intercept (baseline log odds)

⍰N(0, 5^2^)

β_1_, β_2_, β_3_ Fixed effects for individual covariables (age, sex, being in poorest tertile)

⍰N(0, 2.5^2^)

β_4_, β_5_ Fixed effects for neighbourhood covariables (HIV prevalence, population density)

⍰N(0, 2.5^2^)

u_neighbourhood[i]_ Neighbourhood-level random intercept

⍰N(0, σ^2^ _neighbourhood_)

### Model summary

|  | Baseline model | Extended model |
| --- | --- | --- |
| **Age (odds ratio, 95% CrI)** | 1.03 (0.87-1.22) | 1.02 (0.85-1.20) |
| **Sex (female) (odds ratio, 95% CrI)** | 1.39 (0.92-2.09) | 1.36 (0.89-2.06) |
| **Poorest third (odds ratio, 95% CrI)** | NA | 1.88 (1.08-3.01) |
| **HIV prevalence (odds ratio, 95% CrI)** | NA | 1.08 (0.93-1.25) |
| **Population density (odds ratio, 95% CrI)** | NA | 0.84 (0.67-1.04) |
| **Between-neighbourhood variability (Intercept sd)** | 1.76 (1.27-2.55) | 1.55 (1.06-2.33) |
| **Intraclass correlation coefficient (estimate, 95% CrI)** | 0.087 (0.017-0.21) | 0.055 (0.00-0.18) |
| **LOOIC** | 743.03 | 741.55 |
| **WAIC** | 742.88 | 741.39 |
| **Moran's I statistic (estimate, p value)** | 0.03, p = 0.335 | 0.04, p = 0.309 |

sd: standard deviation. LOOIC: Leave-One-Out Information Criterion. WAIC: Widely-Aplicable Information Criterion

