## Supplementary material for "Heterogeneity in *Mycobacterium tuberculosis* immunoreactivity in young children in Blantyre, Malawi: a community-based survey": Ethics approval

UNIVERSITY OF MALAWI

### CERTIFICATE OF ETHICS APPROVAL

This is to certify that the College of Medicine Research and Ethics Committee (COMREC) has reviewed and approved a study entitled:

P.04/22/3611 - Tuberculosis Immunoreactivity Surveillance in Malawi (Timasamala) version 1.0 by Dr Hannah Rickman

*On 09-Jun-22*

*As you proceed with the implementation of your study, we would like you to adhere to international ethical guidelines, national guidelines and all requirements by COMREC some of which are indicated on the next page for your study*

09-Jun-22

**Prof. E. Umar -Chairperson (COMREC)**

Date

Approved by
College of Medicine
09-Jun-2022
(COMREC)
Research and Ethics Committee
