## Supplementary material for "Heterogeneity in *Mycobacterium tuberculosis* immunoreactivity in young children in Blantyre, Malawi: a community-based survey": Ethics approval 2

**London School of Hygiene & Tropical Medicine**
Keppel Street, London WC1E 7HT
United Kingdom
Switchboard: +44 (0)20 7636 8636

**www.lshtm.ac.uk**

**Observational / Interventions Research Ethics Committee**

Dr Hannah Rickman
LSHTM

3 August 2022

Dear Dr Hannah Rickman

**Study Title:** Timasamala - TB Immunoreactivity for Surveillance in Malawi

**LSHTM Ethics Ref:** 27744

Thank you for responding to the Observational Committee's request for further information on the above research and submitting revised documentation.

The further information has been considered on behalf of the Committee by the Chair.

**Confirmation of ethical opinion**

On behalf of the Committee, I am pleased to confirm a favourable ethical opinion for the above research on the basis described in the application form, protocol and supporting documentation as revised, subject to the conditions specified below.

**Conditions of the favourable opinion**

Approval is dependent on local ethical approval having been received, where relevant.

**Approved documents**

The final list of documents reviewed and approved by the Committee is as follows:

| Document Type | File Name | Date | Version |
|---|---|---|---|
| Other | GCP_refresher | 12/05/2020 | 1 |
| Protocol / Proposal | 5a. Translated_CRF_proxymeans | 04/04/2022 | 1 |
| Protocol / Proposal | 5b. Timasamala_interviewguide | 04/04/2022 | 1 |
| Investigator CV | 12a. Hannah Rickman CV | 11/04/2022 | 1 |
| Investigator CV | 12b. Mphatso Phiri CV | 11/04/2022 | 1 |
| Investigator CV | 12c. Helena Feasey CV | 11/04/2022 | 1 |
| Investigator CV | 12d. McEwen Khundi CV | 11/04/2022 | 1 |
| Investigator CV | 12e. Marriott Nliwasa CV | 11/04/2022 | 1 |
| Investigator CV | 12f. Katherine Fielding CV | 11/04/2022 | 1 |
| Investigator CV | 12g. Liz Corbett CV | 11/04/2022 | 1 |
| Investigator CV | 12h. Peter MacPherson CV | 11/04/2022 | 1 |
| Other | Research_Ethics_certificate | 11/04/2022 | 1 |
| Protocol / Proposal | 220531_Timasamala_CRFs | 30/05/2022 | 2 |
| Investigator CV | 12i. Robina Semphere CV | 30/05/2022 | 1 |
| Local Approval | Timasamala_COMREC_cert | 09/06/2022 | 1 |
| Information Sheet | 220801_Timasamala_PILconsent_communityadults_v4_clean | 01/08/2022 | 4.0 |
| Information Sheet | 220801_Timasamala_PILconsent_communityadults_v4_tracked | 01/08/2022 | 4.0 |
| Information Sheet | 220801_Timasamala_PILconsent_communitychildren_v4_clean | 01/08/2022 | 4.0 |
| Information Sheet | 220801_Timasamala_PILconsent_communitychildren_v4_tracked | 01/08/2022 | 4.0 |
| Information Sheet | 220801_Timasamala_PILconsent_PHC_v4_clean | 01/08/2022 | 4.0 |

| | | | |
|---|---|---|---|
| Information Sheet | 220801_Timasamala_PILconsent_PHC_v4_tracked | 01/08/2022 | 4.0 |
| Protocol / Proposal | 220801_Timasamala_protocol_v4_clean | 01/08/2022 | 4.0 |
| Protocol / Proposal | 220801_Timasamala_protocol_v4_tracked | 01/08/2022 | 4.0 |
| Covering Letter | 220801_Timasamala_LSHTMethicsresponse2 | 01/08/2022 | 2 |

**After ethical review**

The Chief Investigator (CI) or delegate is responsible for informing the ethics committee of any subsequent changes to the application. These must be submitted to the Committee for review using an Amendment form. Amendments must not be initiated before receipt of written favourable opinion from the committee.

The CI or delegate is also required to notify the ethics committee of any protocol violations and/or Suspected Unexpected Serious Adverse Reactions (SUSARs) which occur during the project by submitting a Serious Adverse Event form.

An annual report should be submitted to the committee using an Annual Report form on the anniversary of the approval of the study during the lifetime of the study.

At the end of the study, the CI or delegate must notify the committee using an End of Study form.

All aforementioned forms are available on the ethics online applications website and can only be submitted to the committee via the website at: http://leo.lshtm.ac.uk

Additional information is available at: www.lshtm.ac.uk/ethics

Yours sincerely,

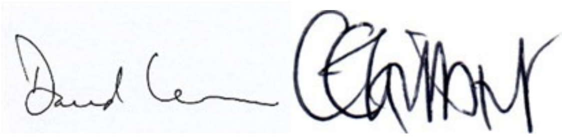

**Professor David Leon and Professor Clare Gilbert**
**Co-Chairs**


http://www.lshtm.ac.uk/ethics/
